# Genomic epidemiology of ESBL-producing *Escherichia coli* and *Klebsiella pneumoniae* across the human-animal-environment interface in peri-urban pig farms in Yaounde, Cameroon

**DOI:** 10.64898/2026.03.16.26348538

**Authors:** Germanie Delaisie Abomo, Gabriel Cedric Bessala, Isaac Dah, Michelle M. C. Buckner, Jan-Ulrich Kreft, Blaise Pascal Bougnom

## Abstract

**Background:** Livestock production systems in peri-urban areas are associated with high levels of interaction between humans, animals, and the environment, which may contribute to the dissemination of antimicrobial resistant bacteria. However, genomic characterization of resistant bacteria in the interconnected systems of humans, animals, and the environment in low- and middle-income countries like Cameroon is very limited.

**Methods:** This study was undertaken to investigate the ESBL-producing *E. coli* and *K. pneumoniae* in the peri-urban pig production systems in Yaounde, Cameroon, through the application of the One Health genomic approach. A total of 338 samples were collected from humans, pigs, and the environment. Enterobacterales were isolated using standard microbiological procedures, followed by antimicrobial susceptibility testing of the isolated bacteria using the Kirby-Bauer disk diffusion method based on the EUCAST breakpoints. Ten multidrug-resistant Enterobacterales with similar resistance profiles were sequenced to identify their sequence types, resistance determinants, plasmid replicons, and virulence determinants.

**Results:** Enterobacterales were found in 187 samples, comprising 38 human, 98 pig, and 51 environmental samples. *E. coli* (166 isolates) was the most prevalent species, followed by *K. pneumoniae* (100 isolates). Whole-genome sequencing revealed eight *E. coli* and two *K. quasipneumoniae* isolates from human, pig, wastewater, and farm environmental samples. The *E. coli* isolates represented seven sequence types, including the globally successful ST410 lineage. Notably, *E. coli* ST3580 was found in human and environmental samples from the Afanoyoa farm in different sampling months, while *K. quasipneumoniae* ST1535 was found in human and pig samples from the Etoudi farm in different months. All genomes encoded ESBL genes, with *bla*_CTX-M-15_ being the most prevalent, accompanied by other resistance genes to various antibiotic classes and several plasmid incompatibility groups.

**Conclusions:** These results show the circulation of genetically diverse ESBL-producing *E. coli* and *K. pneumoniae* in human, animal, and environmental reservoirs in peri-urban pig farming systems and the potential for cross-reservoir persistence of particular lineages. Improved One Health antimicrobial resistance surveillance and stewardship are critical to address antimicrobial resistance in rapidly urbanizing environments.

## 1. INTRODUCTION

Antimicrobial resistance (AMR) is one of the most serious global health issues of the 21st century. AMR jeopardizes our ability to effectively prevent and treat infectious diseases in both humans and animals. The emergence and spread of antibiotic-resistant bacteria (ARB) are mainly consequent to the overuse and misuse of antibiotics in human and animal medicine, agriculture and animal farming. This issue is further intensified by environmental contamination and poor implementation of infection prevention and control (IPC) (Laxminarayan et al., 2013; O’Neill, 2016; Van Boeckel et al., 2015). sub-Saharan Africa suffers heavily from AMR as it has limited legislations, poor implementation of water, sanitation and hygiene (WASH), and weak surveillance systems (Leopold et al., 2014; Okeke & Edelman, 2001).

Among pathogens of concerns are extended-spectrum β-lactamase (ESBL) producing Enterobacterales, particularly ESBL-*Escherichia coli* and *Klebsiella* spp which are among critical pathogens according to the World Health Organization (WHO). They are resistant to third-generation cephalosporins and are associated with severe clinical outcomes. These pathognes can cause community-acquired and healthcare-associated infections, including urinary tract infections, bloodstream infections, and diarrhea, with morbidity, mortality, longer hospital stays, and increased costs to the health care system as the outcomes (Paterson & Bonomo, 2005; Pitout & Laupland, 2008; Schwaber & Carmeli, 2007).

Beyond clinical settings, livestock production systems have increasingly been identified as significant sources and amplifiers of AMR. The extensive application of antimicrobials in animals for food production leads to the selection persistence, and spread of ARB and antibiotic-resistant genes (ARGs). These ARB might later be transmitted to humans through direct contact with animals, the food chain, or via environmental routes like contaminated soil, water, and agricultural waste (Marshall & Levy, 2011; Robinson et al., 2016; Van Boeckel et al., 2019; Woolhouse et al., 2015). Production of pigs is especially significant in this regard, since not only are pigs often given antimicrobials to treat illness, prevent disease, or for growth promotion, but also, they can carry multidrug-resistant ARB including ESBL-*E. coli* and *Klebsiella* spp. (Rhouma et al., 2017; Tang et al., 2017).

Peri-urban livestock farming systems in many low- and middle-income countries (LMICs) are important interfaces where humans, animals, and environmental compartments are in close contact. In open cities like Yaounde, Cameroon, small-scale pig farms are often found in densely populated areas. These farming systems have only limited biosecurity measures in place, poor waste management, and frequent overlap between domestic and farming water sources (Grace et al., 2012; Abomo et al., 2025). This can favor the spread and persistence of ARB across multiple ecological compartments including pigs, farmers, soil, water, and farm surfaces. The disposal of untreated animal waste into the environment surrounding the farm increases the probability of ARB and ARGs to spread beyond the farm and potentially contributing to community-level exposure (Berendonk et al., 2015; Manaia et al., 2016).

Cameroon like many other countries in sub-Saharan Africa faces the burden of AMR. Previous studies have reported high prevalence rates of ESBL-producing *E. coli* and *Klebsiella* spp. in healthcare settings, highlighting the growing clinical impact of these pathogens (Breurec et al., 2013; Lonchel et al., 2012; Ndzi et al., 2016). However, the contribution of animal production systems including pig farming to investigate genomic epidemiology and dissemination of ESBL-producing Enterobacterales remains understudied.

WGS has recently given new dimensions for the study of bacterial genetic epidemiology and AMR surveillance. WGS enables high-resolution characterization of bacterial genomes, allowing the identification of AMR genes, plasmids, virulence factors, and clonal lineages along with the phylogenetic reconstruction of isolates from various ecological niches (Didelot & Maiden, 2010; Didelot et al., 2017; Köser et al., 2012). Using the One Health approach, WGS is the ideal way to investigate transmission routes, identify cross-reservoir spread, and gain insight into the genetic factors driving the dissemination of AMR at the human-animal-environment interface (McEwen & Collignon, 2018; Robinson et al., 2016).

However, there are still knowledge gaps in assessing AMR genomic on the livestock-human-environment interface in sub-Saharan Africa. Specifically, the One Health genomic approach using WGS to assess the spread and persistence of ESBL-producing Enterobacterales in peri-urban pig farming in Yaounde has not yet been fully implemented. Closing such knowledge gaps is crucial to provide guidance on the development of targeted surveillance of AMR and the formulation of policies to mitigate the spread of AMR in community settings.

In this study, we investigated the circulation of ESBL-producing *E. coli* and *Klebsiella* spp. within peri-urban pig farming systems, using One Health approach. By combining antimicrobial susceptibility testing and WGS of isolates collected from pigs, farmers, and environmental samples, we aimed to (i) identify strains with similar phenotypic resistance, (ii) characterize the resistome, plasmid content, and virulence determinants, (ii) examine their population genomic structure, and (iii) assess evidence for possible cross-reservoir transmission at the human-animal-environment interface.

## 2. MATERIALS AND METHODS

### 2.1 Study design and sampling sites

This study was conducted in Yaoundé, the political capital of Cameroon (3°52′N, 11°31′E), with about 5 million inhabitants. Yaoundé features a tropical wet and dry climate with total rainfall and an average temperature of 2011 mm and 24°C, respectively (Nimpa et al. 2023). Four pig farms located in different administrative divisions of the city were selected for the study (Figure 1); namely Yaoundé I (Etoudi), Yaoundé III (Odza), Yaoundé IV (Afanoyoa), and Yaoundé VII (Nkolbisson) (Figure 1).

**Figure 1.**
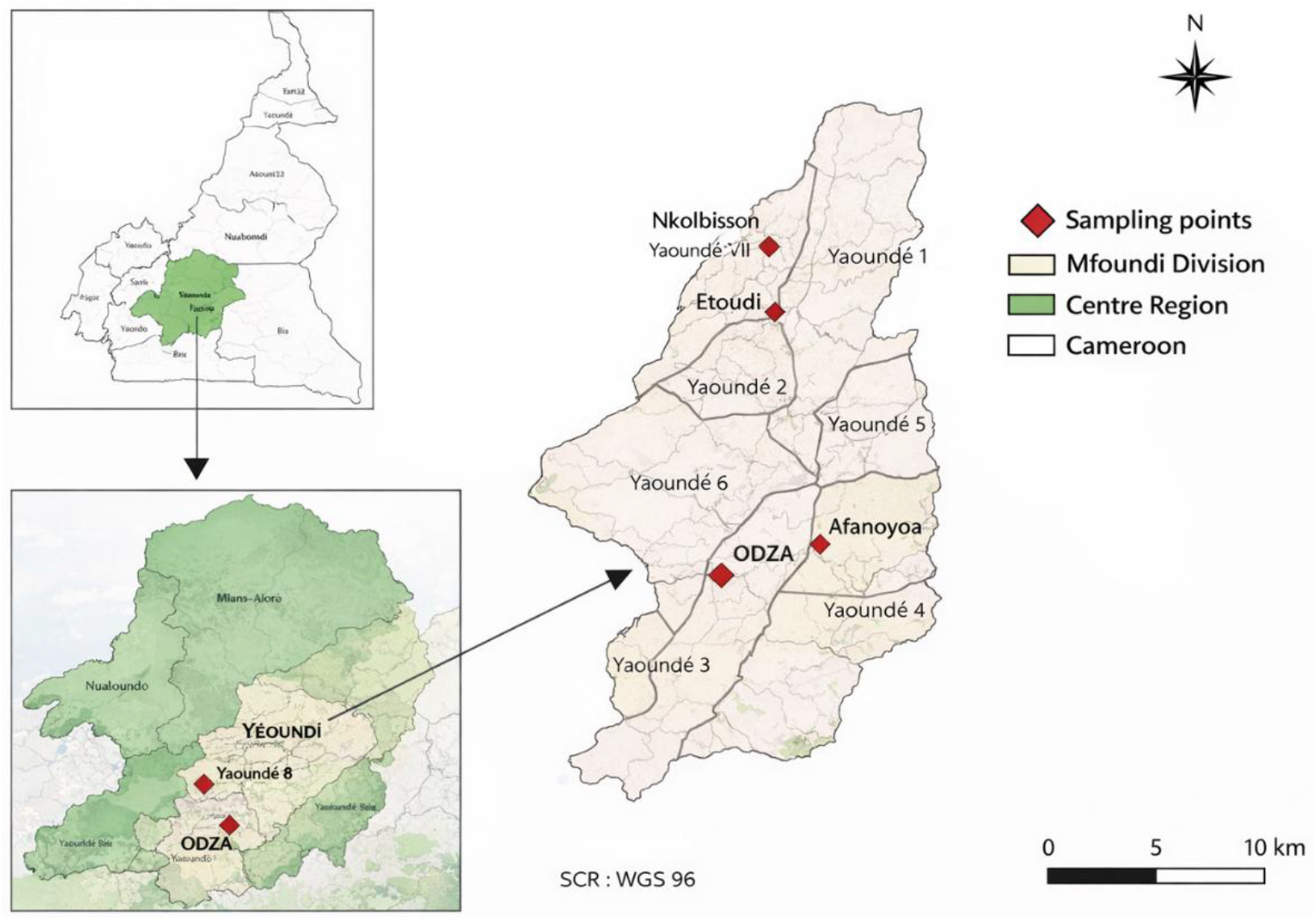
Map of the study area and sampling locations in Yaounde, Cameroon.

The selected farms are peri-urban pig farm production systems commonly found in the city, where livestock production frequently occurs in close proximity to residential areas. They are characterized by intense interactions between humans, animals, and environmental compartments, including soil, water, and farm surfaces, which makes them particularly relevant for the One Health concept investigation approach.

### 2.2 Sampling strategy and sample collection

Sampling was conducted throughout the year 2024. The sample collection was done twice every two months, for a total of eight sampling times. Samples were collected from apparently healthy pigs on family-owned farms. To avoid bias, sick animals or farmers under antibiotic treatment were excluded from the study. In each farm, at each sampling time, samples were collected from the three compartments: pigs, farmers, and the environment. Rectal swabs were obtained from pigs, whereas stool samples were obtained from farmers who were directly involved in animal husbandry activities. Environmental samples included food, soil, water, and surface samples. Approximately 50 g of soil were randomly collected from five different locations within each farm and pooled to get a composite sample representing the farm environment. Three samples were obtained from each water source, including animal drinking water and household water supplies associated with each farm. Additional environmental samples were obtained from farm surfaces frequently in contact with animals, including feeders and drinkers, as well as from footbaths and wastewater collection areas. Samples were collected using sterile containers, transported in ice boxes to the laboratory, and processed within six hours of collection to preserve bacterial viability.

### 2.3 Culture and isolation and of cefotaxin-resistant Enterobacterales

Selective isolation of cefotaxime-resistant Enterobacterales was conducted using MacConkey agar supplemented with cefotaxime (2 mg/L). Pig rectal swabs were suspended in sterile peptone water and vortexed before 0.1 mL of the suspension was plated onto the selective medium. For human samples, approximately 10 mg of stool was suspended in sterile saline, homogenized, and 0.1 mL of the suspension was plated onto MacConkey agar containing cefotaxime. Soil samples were prepared by suspending 10 g of soil in 90 mL of sterile saline, vortexing the mixture, and plating 0.1 mL of the suspension onto the selective agar. For water samples, 0.1 mL of each sample was directly inoculated onto MacConkey agar supplemented with cefotaxime. All plates were incubated at 37 °C for 24 h, after which colonies exhibiting morphological characteristics consistent with Enterobacterales were selected for further analysis.

### 2.4 Bacterial identification

The presumptive colonies of *E. coli* and *Klebsiella* spp. were subcultured on nutrient agar media and incubated at 37 °C for 24 h to obtain pure cultures. Biochemical tests like Kligler Iron Agar, Simmons citrate test, urease-indole test, along with colony characteristics and Gram staining, were used for preliminary identification (Cappuccino & Welsh, 2017). Eosin Methylene Blue agar for *E. coli* and Hektoen agar for *K. pneumoniae*, and the API 20E kit (bioMérieux, France) were used for further identification. The pure cultures were preserved in Brain Heart Infusion broth containing 20% glycerol at -20 °C for further phenotypic and genomic analysis.

### 2.5 Phenotypic detection of ESBL production

Phenotypic detection of ESBL production by *E. coli* and *K. pneumoniae* isolates was conducted using the double-disc synergy test (DDST) (EUCAST, 2024). Suspensions of the bacteria corresponding to the 0.5 McFarland turbidity standard were prepared and inoculated onto Mueller-Hinton agar plates. An amoxicillin/clavulanic acid disc was placed at the center of the inoculated plates, whereas the cefotaxime, ceftazidime, cefixime, and aztreonam discs were placed 20-30 mm away from the center disc. The plates were incubated at 37°C for 18-24 hours, after which the bacteria showing a characteristic increase of the inhibition zone around the amoxicillin/clavulanic acid disc were considered as ESBL producers (EUCAST, 2024).

### 2.6 Antimicrobial susceptibility testing

The AST of ESBL *E. coli* and *K. pneumoniae* producers was done using the Kirby-Bauer disk diffusion method on Mueller-Hinton agar plates following EUCAST guidelines (EUCAST, 2024). Suspensions of bacteria were prepared from an overnight culture of each isolate, adjusted to a turbidity of 0.5 McFarland units, and used to inoculate Mueller-Hinton agar plates. Sixteen antimicrobial discs of various classes of antibiotics commonly used in human and veterinary medicine were placed on the inoculated plates. The plates were incubated at 37°C for 18-24 hours, after which the diameters of the inhibition zones were measured and interpreted as susceptible or resistant based on EUCAST breakpoints.

### 2.7 DNA extraction, library preparation, and whole-genome sequencing

Whole genome sequencing was done at MicrobesNG (University of Birmingham, UK), on ten selected ESBL-producing *E. coli* and *K. pneumoniae* isolates, based on their phenotypic resistance after AST. Genomic DNA of isolates was extracted and genomic libraries were constructed using Illumina-compatible library prep kits following the manufacturer’s protocol. The libraries were sequenced using an Illumina HiSeq 2500 platform to generate 250 bp paired-end reads. Sequencing was conducted at a minimum genome coverage depth of 30x.

### 2.8 Bioinformatics and genomic analyses

Quality assessment of sequencing reads was carried out using FastQC v0.11.9 (Andrews, 2010). Subsequently, trimming of low-quality reads and adapters was performed using Trimmomatic v0.39 (Bolger et al., 2014). Finally, high-quality reads were assembled into contigs de novo using SPAdes v3.15.5 (Bankevich et al., 2012). The quality of assemblies was assessed using QUAST v5.2.0 (Gurevich et al., 2013). Contigs with a length of less than 500 bp and an average coverage of less than 10 were removed for better assembly. Identification of species was carried out using Mash software (Ondov et al., 2016) and Kraken2 software (Wood et al., 2019). Multilocus sequence typing (MLST) was performed using mlst software based on the PubMLST scheme (Jolley & Maiden, 2010). Plasmid replicons were identified using PlasmidFinder with thresholds set to ≥95% identity and ≥60% 162 coverage (Carattoli et al., 2014). Identification of antimicrobial resistance genes and virulence factors was performed using ABRicate against ResFinder v4.7.2 (Zankari et al., 2012), CARD v3.3.0 (Jia et al., 2017), ARG-ANNOT v6 (Gupta et al., 2014), and VFDB v6.0 databases, with settings of ≥90% 164 identity and ≥90% coverage (Chen et al., 2016). For phylogenomic analysis, genome assemblies were annotated using Prokka v1.14.6 (Seemann, 2014). The pangenome analysis was done using Roary v3.13.0 (Page et al., 2015). The results were used to construct a maximum likelihood phylogenetic tree using IQ-TREE v2.1.4 (Nguyen et al., 2015). The best nucleotide substitution model was automatically selected using ModelFinder (Kalyaanamoorthy et al., 2017). The branch support was done using ultrafast bootstrap analysis with 1,000 replicates (Hoang et al., 2018). The phylogenetic trees were visualized and annotated using iTOL v6 (Letunic & Bork, 2021).

### 2.9 Availability of data and materials

The assembled genomes of the 10 isolates have been deposited in the public database (NCBI GenBank database) under BioProject no. PRNJAXXXXX

### 2.10 Ethical considerations

Ethical clearance was sought from the Centre Region Ethics Committee (0125-/CRERSHC/2024) and approval from the Ministry of Livestock, Fisheries and Animal Industries (101/L/DREPIA-CE/DD-MFDI). Informed consent was sought from all the respondents after explaining the purpose of the research.

## 3. RESULTS

In total, 338 samples were collected during the study period, including 168 rectal swabs from pigs, 128 environmental samples, and 42 stool samples from farmers. ESBL-*E. coli* was isolated from 166 samples, including 32 human, 87 pig, and 47 environmental samples. ESBL-*K. pneumoniae* was isolated from 100 samples, including 29 human, 41 pig samples, and 30 environmental samples.

### 3.1 Population structure and sequence types

From the ESBL-*E. coli* and *K. pneumoniae* isolates recovered during culture screening, ten ESBL-producing isolates were selected for WGS, based on their similar phenotypic resistance profiles observed after AST. These included three *E. coli* from humans, three *E. coli* from pigs, two *E coli* from environmental samples (one from wastewater sample and from footbath sample), one *K. pneumoniae* from a human, and one *K. pneumoniae* from a pig. These isolates originated from the four pig farms, and were isolated between January and October 2024.

Genome-based identification after MLST revealed that the eight *E. coli* isolates were a genetically diverse population, with seven distinct sequence types (STs): ST3580, ST4389, ST410, ST2705, ST3716, ST4684, and ST3274. Two isolates belonged to ST3580, both sampled from the Afanoyoa farm, with one recovered from a human clinical sample and the other from an environmental footbath sample. The two *K. quasipneumoniae* isolates belonged to ST1535 and were isolated from the Etoudi farm, one from a pig sample and the other from a human clinical sample. The characteristics of the isolates, including species, sequence type, source, farm location, and sampling period, are presented in Table 1.

**Table 1.** Characteristics of ESBL *E. coli* and *K. quasipneumoniae* isolates recovered from human, animal, and environmental sources in peri-urban farms of Yaoundé, Cameroon.

| Isolates | Source | Sample Type | Farm | Sampling Month |
| --- | --- | --- | --- | --- |
| *E. coli* ST3580 | Human | Clinical | Afanoyoa | June |
| *E. coli* ST3580 | Environment | Footbath | Afanoyoa | January |
| *E. coli* ST4389 | Animal | Pig | Afanoyoa | January |
| *E. coli* ST410 | Human | Clinical | Odza | January |
| *E. coli* ST2705 | Animal | Pig | Odza | October |
| *E. coli* ST3716 | Environment | Wastewater | Nkolbisson | September |
| *E. coli* ST4684 | Human | Clinical | Nkolbisson | August |
| *E. coli* ST3274 | Animal | Pig | Nkolbisson | January |
| *K. quasipneumoniae* ST1535 | Animal | Pig | Etoudi | January |
| *K. quasipneumoniae* ST1535 | Human | Clinical | Etoudi | March |

Footnote: Isolates were obtained from peri-urban farming systems representing the human-animal-environment interface. Sequence types (STs) were assigned using multilocus sequence typing (MLST).

### 3.2 Phylogenetic relationships

Core-genome phylogenetic analysis revealed substantial genetic diversity among *E. coli* isolates, indicating a polyphyletic population structure. The different ESBL-*E. coli* isolates were distributed across several distinct phylogenetic lineages. The two ST3580 isolates from Afanoyoa formed a monophyletic clade with minimal genetic divergence, indicating a close genomic relationship between the human and environmental isolates. Other *E. coli* lineages, including ST410, ST3716, ST4389, ST2705, ST3274, and ST4684, were phylogenetically distinct and dispersed across the tree (Figure 2).

**Figure 2.**
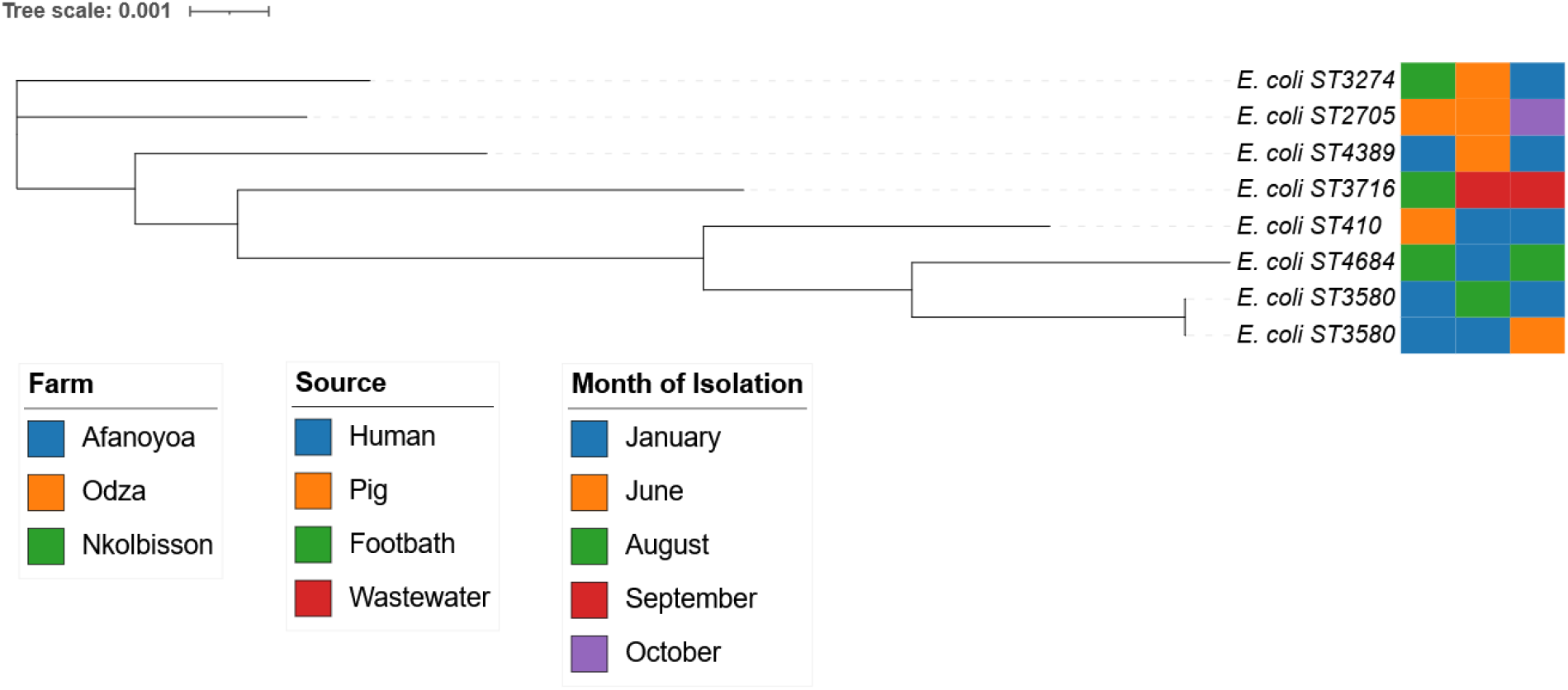
Core-genome phylogenetic tree of the selected eight ESBL-producing *E. coli* isolates recovered from human, pig, and environmental sources in peri-urban pig farms in Yaounde, Cameroon.

### 3.3 Plasmid replicons, antimicrobial resistance genes and virulence factors

The isolates carried diverse plasmid replicons. Various plasmid replicon incompatibility groups were found, including IncF family plasmids (IncFIA, IncFIB, IncFII), IncHI1B, IncHI2/IncHI2A, IncX1, IncP1, and p0111. Small mobilizable Col-type plasmids as well as Col156, Col440I, and Col (MG828) were also found. *E. coli* ST410 harbors IncFIA, IncFIB, and IncFII plasmid replicons. The two *K. quasipneumoniae* ST1535 isolates and the sewage *E. coli* ST3716 isolate possessed IncHI1B plasmid replicon. *E. coli* ST3274 from pig at the Nkolbisson farm had IncHI2-related plasmids, whereas *E. coli* ST3580 isolate from human at the Afanoya farm was lacking any plasmid replicons. Visual representation of the distribution of plasmid replicons among isolates is reported in figure 3.

**Figure 3.**
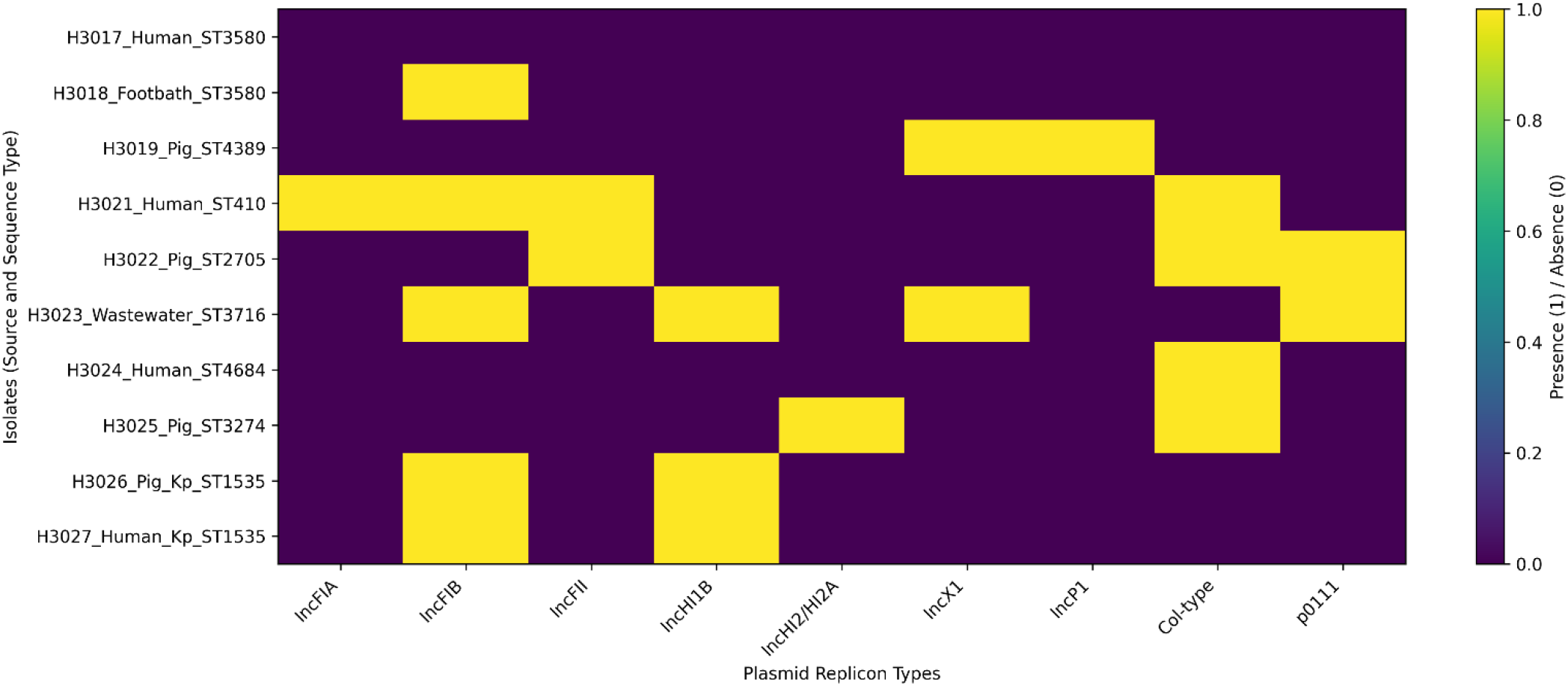
Plasmid replicon distribution heatmap showing the presence and absence of major plasmid incompatibility groups among ESBL *E. coli* and *K. quasipneumoniae* isolates from human, pig, wastewater, and environmental farm sources. The figure highlights the heterogeneous plasmid content across isolates, including IncF (IncFIA, IncFIB, IncFII), IncHI1B, IncHI2/HI2A, IncX1, IncP1, Col-type plasmids, and p0111 replicons.

Genes encoding resistance to various antibiotic families were found in all isolates. ESBL genes were found in the genome of all isolates, and *bla*_CTX-M-15_ was found in all human, animal, and environmental isolates. Other ESBL genes were *bla*_CTX-M-55_, *bla*_TEM-1_, and *bla*_OKP-B-45_. Other resistance genes included *qnrS1* (quinolone resistance), *aadA5, aph(3′)-Ia, aph(3″)-Ib*, and *aph(6)-Id* (aminoglycoside resistance), and *tet(A)* (tetracycline resistance). Fosfomycin resistance genes *fosA* and *fosA7*.*5* were found in both *E. coli* and *Klebsiella* isolates. Other resistance genes included *qacEΔ1* and *fusB* (Figure 4).

**Figure 4.**
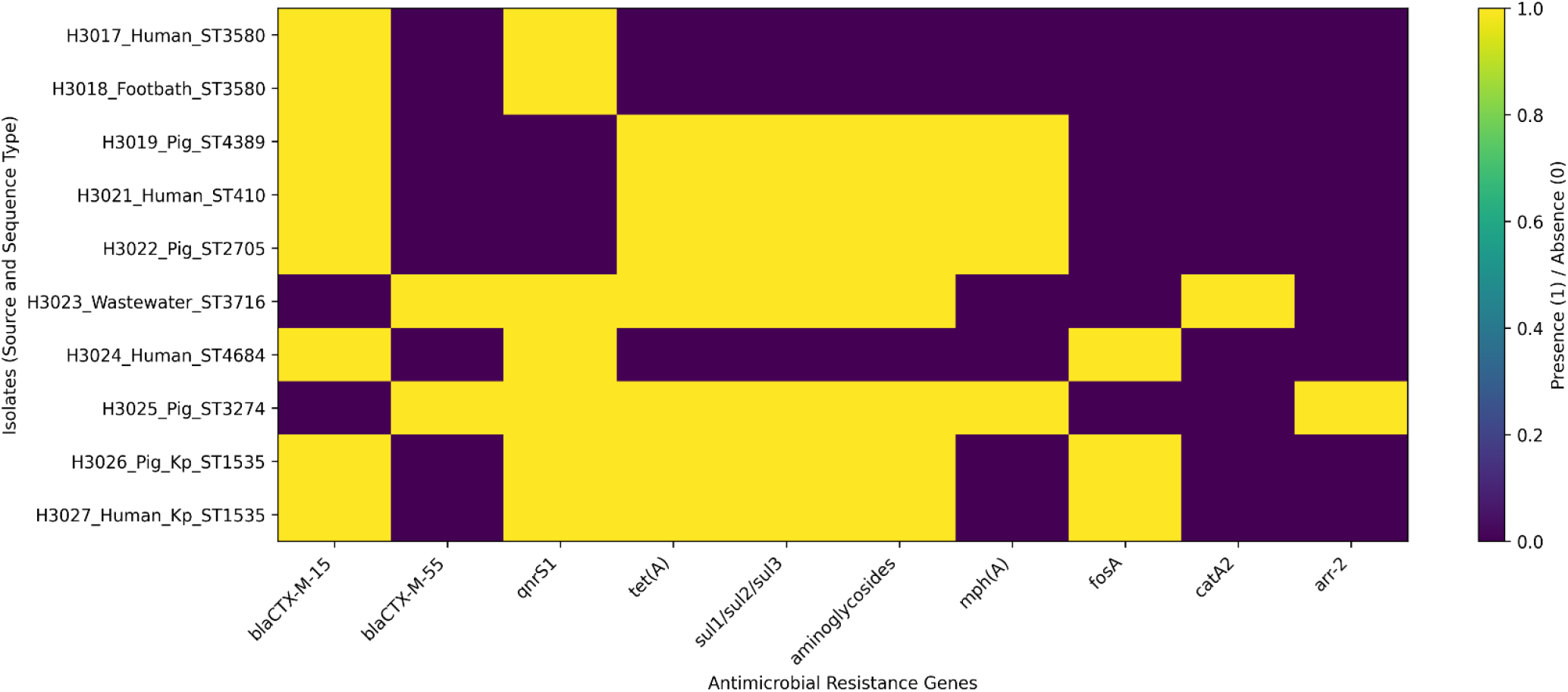
Resistome heatmap showing the distribution of key antimicrobial resistance genes among Enterobacterales isolates from human, pig, wastewater, and environmental farm sources. The heatmap illustrates the presence (1) and absence (0) of major resistance determinants, including ESBL genes (*bla*_CTX-M-15_, *bla*_CTX-M-55_), plasmid-mediated quinolone resistance (*qnrS1*), tetracycline resistance (*tet(A)*), sulfonamide resistance (*sul* genes), aminoglycoside resistance genes, macrolide resistance (*mph(A)*), fosfomycin resistance (*fosA*), chloramphenicol resistance (*catA2*), and rifampicin resistance (*arr-2*).

**Figure 5.**
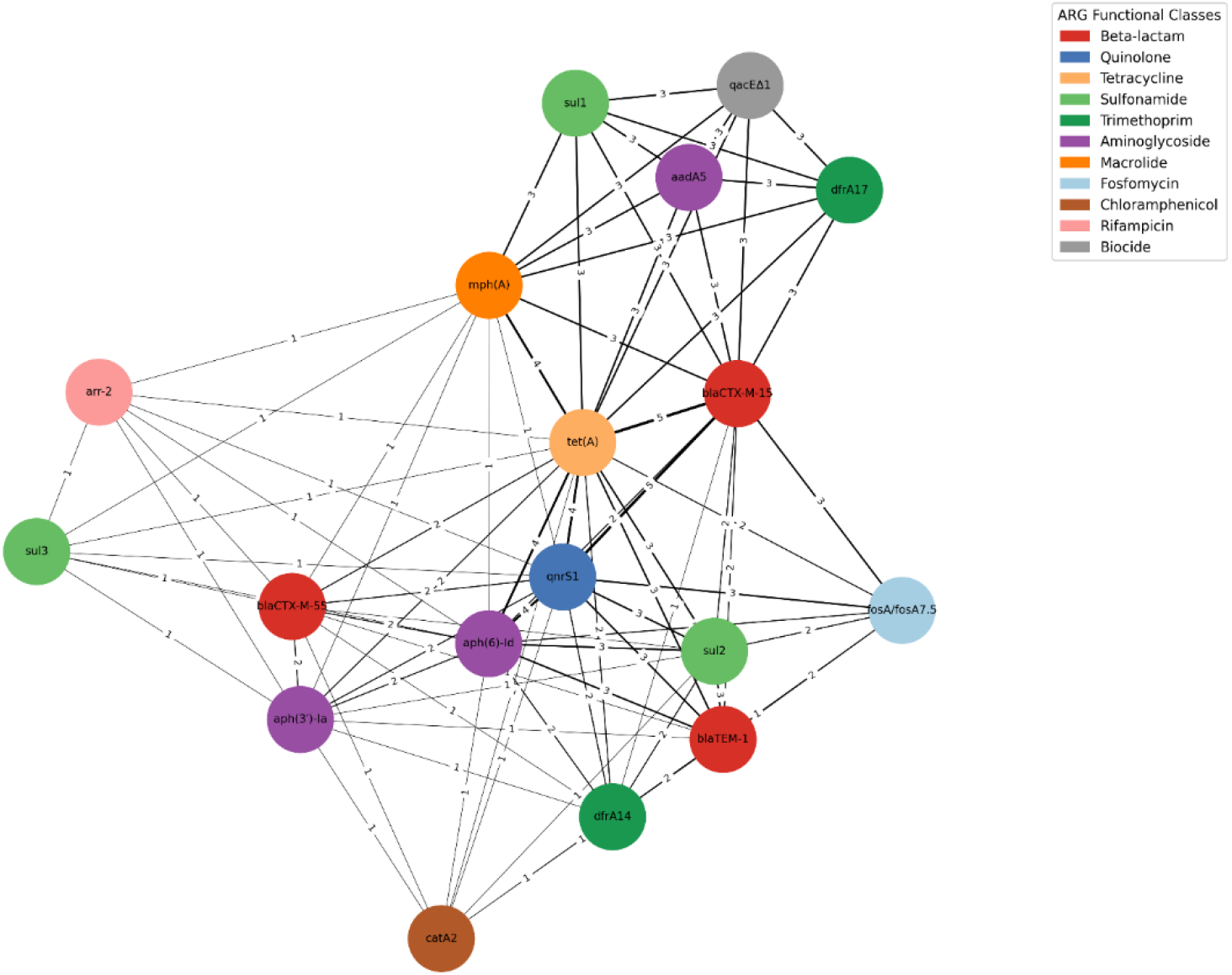
Antimicrobial resistance (AMR) gene co-occurrence network among ESBL *E. coli* and *K. quasipneumoniae* isolates recovered from human, animal, wastewater, and environmental farm sources. Nodes represent individual resistance genes, and edges indicate co-occurrence within the same isolate. Edge weights correspond to the frequency of co-detection across isolates, highlighting dominant resistance clusters including *bla*_CTX-M_ variants, *qnrS1, tet(A), sul* genes, aminoglycoside resistance genes, and *mph(A)*.

The associations between the resistance genes were further investigated using a co-occurrence network analysis, which showed that ESBL genes are often found together with resistance genes to other antimicrobial agents, creating a multidrug resistance gene cluster in the isolates (Figure 4).

All isolates carried multiple virulence-associated genes. Virulence associated to adhesion and fimbrial genes were detected in several isolates, including *fim, ecp, csg, stg*, and *cfa*. Virulence associated to iron acquisition systems included *ent, fep, fes, sit, iuc/iut*, and *ybt/fyuA*, those associated with bacterial motility and chemotaxis, including *flg, fli, flh*, and *che*, were also present in several isolates. Genes associated with type III and type VI secretion systems such as *esp, espX, espL, clpV, hcp*, and *vgrG*, as well as those associated with the general secretion pathway (*gsp* and *epr*) were identified in many isolates (Figure 6).

**Figure 6.**
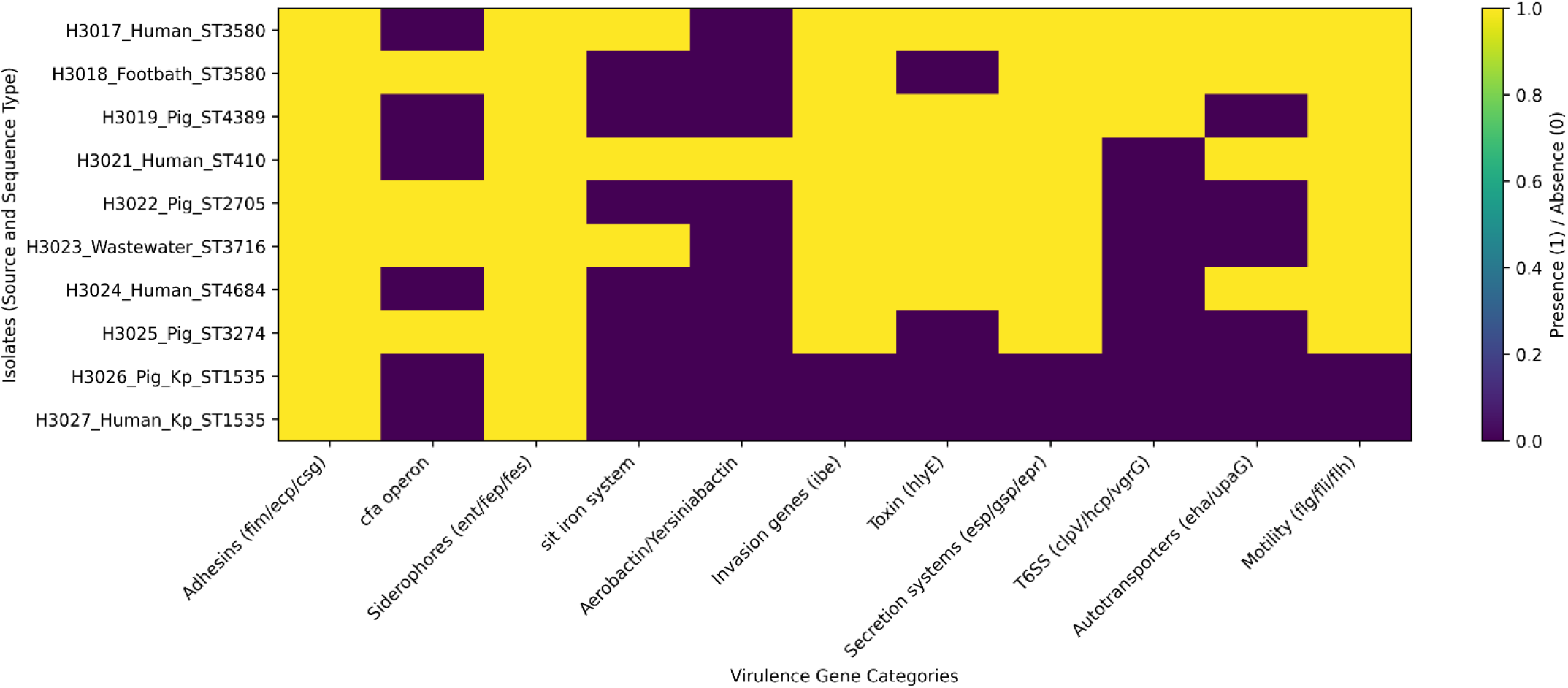
Virulence gene profile heatmap showing the distribution of major virulence-associated gene categories among ESBL *E. coli* and *K. quasipneumoniae* isolates from human, pig, wastewater, and environmental farm sources. The heatmap illustrates the presence (1) and absence (0) of key virulence determinants, including adhesion systems (*fim, ecp, csg*), colonization-associated cfa operon, siderophore and iron acquisition systems (*ent, fep, fes, sit*), aerobactin/yersiniabactin, invasion genes (*ibe*), toxin gene (*hlyE*), secretion systems (*esp, gsp, epr*), type VI secretion components (*clpV/hcp/vgrG*), autotransporters (*eha, upaG*), and motility clusters (*flg, fli, flh*).

### 3.4 Phenotypic antimicrobial susceptibility profiles

All the isolates showed multidrug-resistant phenotypes, expressing resistance to various classes of antibiotics used in human and veterinary medicine. Most of the isolates showed consistent resistance to various β-lactam antibiotics, i.e., ampicillin, amoxicillin, cefixime, cefotaxime, ceftazidime, cefepime, and aztreonam. Resistance to fluoroquinolones, tetracyclines, and sulfamethoxazole/trimethoprim was also expressed by the isolates. Some *E. coli* isolates, i.e., ST3274, ST2705, ST4389, ST3716, and ST410, showed resistance to ciprofloxacin, tetracycline, and sulfamethoxazole/trimethoprim. However, variable susceptibility of the isolates to aminoglycosides and β-lactam/β-lactamase inhibitor combinations was noted. Some of the isolates showed susceptibility to amikacin (07) and gentamicin (05). Similarly, susceptibility to piperacillin/tazobactam (06) and cefoxitin (08) was noted for some of the isolates. Carbapenem susceptibility was relatively preserved with most isolates remaining susceptible to imipenem (06). However, *E. coli* isolate ST410, ST4389, and ST4684 were resistant to imipenem. The pig isolate *K. quasipneumoniae* ST1535 from Etoudi farm was resistant to all tested antibiotics (Table S1).

## 4. DISCUSSION

In this work, we investigated the spread and features of ESBL-producing E. coli and K. pneumoniae in human, animal, and environmental reservoirs in peri-urban pig farming settings of Yaounde, Cameroon. Using AST and WGS, selected isolates were assessed for identity, resistome, plasmid content, and virulence attributes. The outcomes revealed the coexistence of diverse genetic lineages that harbor multiple AMRl resistance factors in interconnected One Health compartments and over multiple sampling periods.

ESBL producing Enterobacterales were present in human, pig, and environmental samples, which supports the idea that these bacteria exist in several reservoirs in peri-urban farming environments. They were more prevalent in human samples, followed by pig and environmental samples. E. coli was the most commonly isolated from the different reservoirs. The extensive presence of ESBL-producing *E. coli* and *K. pneumoniae* indicate active circulation of these bugs between humans, animals and the environment in peri-urban livestock systems. Similar observations have been reported in previous One Health studies focusing AMR in livestock production systems of LMICs, where biosecurity measures are limited and poor waste management can lead to the spread of resistant bacteria among different reservoirs (Robinson et al., 2016; McEwen & Collignon, 2018).

The multidrug-resistant isolates were isolated at several time points during the whole sampling period, for example in January, March, June, August, September, and October. This temporal distribution suggests that ESBL-producing *E. coli* and *K. pneumoniae* circulated during the study period rather than the emergence of a single outbreak.

For example, the detection of the ST3580 lineage in both human and environmental footbath from Afanoyoa farm, as well as *K. quasipneumoniae* ST1535 from Etoudi farm at different sampling times, might indicate the continued presence of particular bacterial lineages in the farm environment. Such presence could be consequent to environmental contamination, insufficient cleaning, or constant contact between animals, humans, and farm surfaces.

WGS revealed high level of genetic variation with seven different STs among eight *E. coli* genomes. The core-genome phylogeny shows a polyphyletic structure, which means the resistant population consisted of multiple unrelated lineages instead of being dominated by one single epidemic clone.

Among the detected ST, the globally disseminated lineage *E. coli* ST410 from a human isolate is most remarkable. This lineage has been reported often both in clinical and environmental settings and is usually linked with ESBL production and multidrug resistance (Nicolas-Chanoine et al., 2014; Tchesnokova et al., 2019). Its occurrence in the peri-urban farming environment may indicate the environmental, community, and clinically relevant bacterial population overlap. On the other hand, the close phylogenetic relationship between the two ST3580 isolates isolated from human and environmental samples from the same farm indicates possible local transmission and environmental persistence of this lineage.

WGS unveiled the presence of a wide range of AMR genes in the various isolates. All isolates carried ESBL coding genes, among which *bla*_CTX-M-15_ was, by far, the most frequently detected one. This is illustrated by the resistome heatmap, where the different AMR genes in the isolates are distributed, and which clearly reveals the presence of several resistance genes within a single genome.

The dominance of *bla*_CTX-M-15_ aligns with worldwide studies identifying this gene as the most dominant beta-lactamase gene among Enterobacterales (Storberg, 2014). In Africa, *bla*_CTX-M-15_ has been frequently documented not only in clinical samples but also in samples of animal and environmental origin, thus indicating its wide spread across different reservoirs (Ramatla et al., 2023). Apart from beta-lactamase genes, a number of isolates carried genes conferring resistance to other AMR classes, e.g., quinolones (*qnrS1*), tetracyclines (*tet(A)*), aminoglycosides (*aadA5* and *aph* genes), sulfonamides (*sul* genes) and trimethoprim (*dfrA* genes). The coexistence of these genes in the same isolates indicates the MDR nature of the bacteria populations that are present in these farm environments.

Plasmid replicons characterization unveiled the existence of multiple plasmid replicons incompatibility groups such as IncF, IncHI1B, IncHI2, IncX1, and IncP1. These plasmids are known to be key players in the horizontal gene transfer (HGT) of AMR genes across Enterobacterales owing to their ability to carry multiple resistance determinants and move between bacterial hosts (Carattoli, 2013; Partridge et al., 2018).

The presence of IncF plasmids in the ST410 isolate is consistent with previous reports associated this plasmid family to ESBL-producing *E. coli*. Similarly, IncHI plasmids detected in environmental and animal isolates have been frequently linked with the dissemination of MDR genes in wastewater and farm settings. The detection of these plasmids across isolates from different hosts and environmental compartments suggests that HGT contributes significantly to the spread of AMR within peri-urban livestock farming systems.

The isolates analyzed in this study carried a wide range of virulence-associated genes involved in adhesion, iron acquisition, motility, and secretion systems. These virulence factors may contribute to bacterial colonization, survival in host environments, and pathogenic potential. The ST410 isolate exhibited a virulence gene profile consistent with extraintestinal pathogenic *E. coli* (ExPEC) that can cause infections outside the intestinal tract. Previous studies have reported that ST410 lineages can combine both multidrug resistance and virulence traits, thereby increasing their ability to cause human infections (Johnson & Russo, 2018). Virulence genes were also detected in isolates originating from animals and environmental samples, indicating that strains circulating in livestock environments may possess pathogenic potential.

Comparative genomic analysis showed that human, animal, and environmental source isolates shared sequence types, plasmid replicons, AMR genes, and virulence factors. The finding of closely related isolates in different compartments is a strong indicator of the circulation of ESBL-producing *E. coli* and *K. pneumoniae* between hosts and environmental reservoirs in farming systems. Environmental components such as wastewater, farm surfaces, and contaminated soil may serve as intermediate reservoirs through which resistant bacteria are transmitted from animals to humans. Similar transmission pathways have been reported in One Health studies investigating AMR in agricultural settings (Bougnom et al., 2020; Amoah et al., 2021).

The findings of this study highlight the importance of integrated One Health approach to tackle AMR that include human, animal, and environmental compartments. Surveillance systems focusing exclusively on clinical settings may underestimate significant reservoirs of AMR present in livestock production systems and environmental matrices.

In rapidly urbanizing cities such as Yaounde, peri-urban livestock farms may represent critical ecological interfaces where critical antibiotic-resistant bacteria emerge and spread across reservoirs. Strengthening AMR stewardship, infection and prevention control (IPC), improving water, sanitation and hygiene (WASH), and expanding genomic surveillance in livestock production systems will be essential for mitigating the spread of AMR.

## 5. CONCLUSION

This research provides insight into the genomic characterization of multidrug-resistant Enterobacterales from the human, animal, and environmental compartments of peri-urban pig farming in Yaounde, Cameroon. The prevalence of ESBL-producing *E. coli* and *Klebsiella spp*. was high in the different compartments of the peri-urban pig farming system.

The genomic analysis showed that the ESBL-producing isolates belonged to genetically diverse lineages, such as the globally disseminated *E. coli* ST410 and *Klebsiella quasipneumoniae* ST1535. The wide distribution of ESBL genes such as *bla*_CTX-M-15_ and other additional resistance genes confirmed the multidrug-resistant nature of the isolates. The presence of similar ESBL-producing lineages in the different compartments of the peri-urban pig farming system and the presence of transferable plasmids suggest their persistence and possible transmission among the compartments.

The findings of this research suggest that peri-urban livestock farming is an important interface in the dissemination of antimicrobial-resistant bacteria and that antimicrobial stewardship and integrated One Health genomic surveillance of peri-urban pig farming in Yaoundé and other urbanizing regions of Cameroon are critical to mitigate the dissemination of multidrug-resistant Enterobacterales.

## Data Availability

All data produced in the present study are available upon reasonable request to the authors

## References

Abomo, G. D., Bessala, G. C., Dah, I., & Bougnom, B. P. (2025). One Health Surveillance of ESBL-Producing Escherichia coli and Klebsiella pneumoniae in Pig Farms in Yaounde, Cameroon. Journal of Medical Science, Biology, and Chemistry, 2(2), 224–234. 10.69739/jmsbc.v2i2.1070

Amoah, I. D., Kumari, S., Bux, F., & Reddy, P. (2021). The role of wastewater treatment plants in the dissemination of antibiotic resistance in the environment: A review. Science of the Total Environment, 760, 144160. 10.1016/j.scitotenv.2020.144160

Andrews, S. (2010). FastQC: A quality control tool for high throughput sequence data. Babraham Bioinformatics. https://www.bioinformatics.babraham.ac.uk/projects/fastqc/

Bankevich, A., Nurk, S., Antipov, D., Gurevich, A. A., Dvorkin, M., Kulikov, A. S., Lesin, V. M., Nikolenko, S. I., Pham, S., Prjibelski, A. D., Pyshkin, A. V., Sirotkin, A. V., Vyahhi, N., Tesler, G., Alekseyev, M. A., & Pevzner, P. A. (2012). SPAdes: A new genome assembly algorithm and its applications to single-cell sequencing. Journal of Computational Biology, 19(5), 455–477. 10.1089/cmb.2012.0021

Berendonk, T. U., Manaia, C. M., Merlin, C., Fatta-Kassinos, D., Cytryn, E., Walsh, F., Bürgmann, H., Sørum, H., Norström, M., Pons, M. N., Kreuzinger, N., Huovinen, P., Stefani, S., Schwartz, T., Kisand, V., Baquero, F., & Martinez, J. L. (2015). Tackling antibiotic resistance: The environmental framework. Nature Reviews Microbiology, 13(5), 310–317. 10.1038/nrmicro3439

Bolger, A. M., Lohse, M., & Usadel, B. (2014). Trimmomatic: A flexible trimmer for Illumina sequence data. Bioinformatics, 30(15), 2114–2120. 10.1093/bioinformatics/btu170

Bougnom, B. P., Thiele-Bruhn, S., Ricci, V., Zongo, C., Piddock, L. J. V., & Bougnom, B. P. (2020). Raw wastewater irrigation for urban agriculture in Africa increases the abundance of transferable antibiotic resistance genes in soil, including those encoding extended-spectrum β-lactamases. Science of the Total Environment, 698, 134201. 10.1016/j.scitotenv.2019.134201

Breurec, S., Guessennd, N., Timinouni, M., Le, T. A., Cao, V., Ngandjio, A., Randrianirina, F., Thiberge, J. M., Zriouil, S. B., & Andremont, A. (2013). Klebsiella pneumoniae resistant to third-generation cephalosporins in sub-Saharan Africa. Journal of Antimicrobial Chemotherapy, 68(7), 1550–1558. 10.1093/jac/dkt056

Cappuccino, J. G., & Welsh, C. T. (2017). Microbiology: A laboratory manual (11th ed.). Pearson.

Carattoli, A. (2013). Plasmids and the spread of resistance. International Journal of Medical Microbiology, 303(6–7), 298–304. 10.1016/j.ijmm.2013.02.001

Carattoli, A., Zankari, E., García-Fernández, A., Voldby Larsen, M., Lund, O., Villa, L., Møller Aarestrup, F., & Hasman, H. (2014). In silico detection and typing of plasmids using PlasmidFinder and plasmid MLST. Antimicrobial Agents and Chemotherapy, 58(7), 3895–3903.

Chen, L., Yang, J., Yu, J., Yao, Z., Sun, L., Shen, Y., & Jin, Q. (2016). VFDB: A reference database for bacterial virulence factors. Nucleic Acids Research, 44(D1), D694–D697.

Didelot, X., & Maiden, M. C. J. (2010). Impact of recombination on bacterial evolution. Trends in Microbiology, 18(7), 315–322.

Didelot, X., Bowden, R., Wilson, D. J., Peto, T. E. A., & Crook, D. W. (2017). Transforming clinical microbiology with bacterial genome sequencing. Nature Reviews Genetics, 13(9), 601–612.

European Committee on Antimicrobial Susceptibility Testing (EUCAST). (2024). Breakpoint tables for interpretation of MICs and zone diameters. https://www.eucast.org

Grace, D., Randolph, T., Affognon, H., Dramane, D., Diall, O., & Clausen, P. H. (2012). Characterisation of livestock production systems in developing countries. Acta Tropica, 122(1), 62–71.

Gurevich, A., Saveliev, V., Vyahhi, N., & Tesler, G. (2013). QUAST: Quality assessment tool for genome assemblies. Bioinformatics, 29(8), 1072–1075.

Gupta, S. K., Padmanabhan, B. R., Diene, S. M., Lopez-Rojas, R., Kempf, M., Landraud, L., & Rolain, J. M. (2014). ARG-ANNOT, a new bioinformatic tool to discover antibiotic resistance genes in bacterial genomes. Antimicrobial Agents and Chemotherapy, 58(1), 212–220.

Hoang, D. T., Chernomor, O., Von Haeseler, A., Minh, B. Q., & Vinh, L. S. (2018). UFBoot2: Improving the ultrafast bootstrap approximation. Molecular Biology and Evolution, 35(2), 518–522.

Jia, B., Raphenya, A. R., Alcock, B., Waglechner, N., Guo, P., Tsang, K. K., Lago, B. A., Dave, B. M., Pereira, S., Sharma, A. N., Doshi, S., Courtot, M., Lo, R., Williams, L. E., Frye, J. G., Elsayegh, T., Sardar, D., Westman, E. L., Pawlowski, A. C., … McArthur, A. G. (2017). CARD: The comprehensive antibiotic resistance database. Nucleic Acids Research, 45(D1), D566–D573.

Johnson, J. R., & Russo, T. A. (2018). Extraintestinal pathogenic Escherichia coli. Clinical Microbiology Reviews, 31(4), e00088–17.

Jolley, K. A., & Maiden, M. C. J. (2010). BIGSdb: Scalable analysis of bacterial genome variation. BMC Bioinformatics, 11, 595.

Kalyaanamoorthy, S., Minh, B. Q., Wong, T. K. F., Von Haeseler, A., & Jermiin, L. S. (2017). ModelFinder: Fast model selection for accurate phylogenetic estimates. Nature Methods, 14, 587–589.

Köser, C. U., Ellington, M. J., Cartwright, E. J. P., Gillespie, S. H., Brown, N. M., Farrington, M., Holden, M. T., Dougan, G., Bentley, S. D., Parkhill, J., & Peacock, S. J. (2012). Routine use of microbial whole genome sequencing in diagnostic microbiology. PLoS Pathogens, 8(8), e1002824.

Leopold, S. J., Van Leth, F., Tarekegn, H., & Schultsz, C. (2014). Antimicrobial drug resistance among clinically relevant bacterial isolates in sub-Saharan Africa. Journal of Antimicrobial Chemotherapy, 69(9), 2337–2353.

Letunic, I., & Bork, P. (2021). Interactive Tree Of Life (iTOL) v6: An online tool for phylogenetic tree display and annotation. Nucleic Acids Research, 49(W1), W293–W296.

Lonchel, C. M., Meex, C., Gangoué-Piéboji, J., Boreux, R., Assoumou, M. C., Melin, P., & De Mol, P. (2012). Proportion of extended-spectrum β-lactamase producing Enterobacteriaceae in a community setting in Cameroon. BMC Infectious Diseases, 12, 53.

Laxminarayan, R., et al. (2013). Antibiotic resistance-the need for global solutions. The Lancet Infectious Diseases, 13(12), 1057–1098.

Manaia, C. M., Rocha, J., Scaccia, N., Marano, R., Radu, E., Biancullo, F., Cerqueira, F., Fortunato, G., Iakovides, I., Zammit, I., Kampouris, I., Vaz-Moreira, I., Nunes, O. C., & Fatta-Kassinos, D. (2016). Antibiotic resistance in wastewater treatment plants. FEMS Microbiology Reviews, 40(5), 849–880.

Marshall, B. M., & Levy, S. B. (2011). Food animals and antimicrobials. Clinical Microbiology Reviews, 24(4), 718–733.

McEwen, S. A., & Collignon, P. J. (2018). Antimicrobial resistance: A One Health perspective. Microbiology Spectrum, 6(2).

Nguyen, L. T., Schmidt, H. A., Von Haeseler, A., & Minh, B. Q. (2015). IQ-TREE: A fast algorithm for estimating maximum likelihood phylogenies. Molecular Biology and Evolution, 32(1), 268–274.

Nimpa, T. (2023). Analysis of Weather Anomalies to Assess the 2021 Flood Events in Yaounde, Cameroon (Central Africa). American Journal of Climate Change, 12(2), 163–176. 10.4236/ajcc.2023.122014

Nicolas-Chanoine, M. H., Bertrand, X., & Madec, J. Y. (2014). Escherichia coli ST131. Clinical Microbiology Reviews, 27(3), 543–574.

Ondov, B. D., Treangen, T. J., Melsted, P., Mallonee, A. B., Bergman, N. H., Koren, S., & Phillippy, A. M. (2016). Mash: Fast genome distance estimation. Genome Biology, 17, 132.

Page, A. J., Cummins, C. A., Hunt, M., Wong, V. K., Reuter, S., Holden, M. T., Fookes, M., Falush, D., Keane, J. A., & Parkhill, J. (2015). Roary: Rapid prokaryote pan genome analysis. Bioinformatics, 31(22), 3691–3693.

Partridge, S. R., Kwong, S. M., Firth, N., & Jensen, S. O. (2018). Mobile genetic elements associated with antimicrobial resistance. Clinical Microbiology Reviews, 31(4), e00088–17.

Pitout, J. D. D., & Laupland, K. B. (2008). Extended-spectrum β-lactamase-producing Enterobacteriaceae. The Lancet Infectious Diseases, 8(3), 159–166.

Ramatla, T., Ngoma, L., Adetunji, M., Mwanza, M., & Ndou, R. V. (2023). Extended-spectrum β-lactamase producing Enterobacterales in Africa. Frontiers in Microbiology, 14, 1102782.

Robinson, T. P., et al. (2016). Antibiotic resistance is the quintessential One Health issue. Transactions of the Royal Society of Tropical Medicine and Hygiene, 110(7), 377–380.

Storberg, V. (2014). ESBL-producing Enterobacteriaceae in Africa. Infection Ecology & Epidemiology, 4, 20342.

Tang, K. L., et al. (2017). Restricting antibiotic use in food-producing animals. The Lancet Planetary Health, 1(8), e316–e327.

Tchesnokova, V., Radey, M., Chattopadhyay, S., Larson, L., Weaver, J. L., Kisiela, D., Sokurenko, E. V., & Johnson, J. R. (2019). Pandemic fluoroquinolone-resistant Escherichia coli clone ST410. Nature Communications, 10, 3326.

Van Boeckel, T. P., Brower, C., Gilbert, M., Grenfell, B., Levin, S., Robinson, T., Teillant, A., & Laxminarayan, R. (2015). Global trends in antimicrobial use in food animals. PNAS, 112(18), 5649–5654.

Van Boeckel, T. P., Glennon, E. E., Chen, D., Gilbert, M., Robinson, T., Grenfell, B., Levin, S., Bonhoeffer, S., & Laxminarayan, R. (2019). Reducing antimicrobial use in food animals. Science, 357, 1350–1352.

Wood, D. E., Lu, J., & Langmead, B. (2019). Kraken 2: Improved metagenomic analysis. Genome Biology, 20, 257.

Zankari, E., Hasman, H., Cosentino, S., Vestergaard, M., Rasmussen, S., Lund, O., Aarestrup, F. M., & Larsen, M. V. (2012). Identification of antimicrobial resistance genes. Journal of Antimicrobial Chemotherapy, 67(11), 2640–2644.

