## Supplementary Table S1 for "Genomic epidemiology of ESBL-producing *Escherichia coli* and *Klebsiella pneumoniae* across the human-animal-environment interface in peri-urban pig farms in Yaounde, Cameroon"

**Table S1**. Antimicrobial susceptibility profiles of ESBL- producing E. coli and K. quasipneumoniae isolates recovered from humans, animals, and the environment in peri-urban farms of Yaoundé, Cameroon.

| Isolates | Source | Farm | AMP | AMX | CFM | CTX | CAZ | FEP | ATM | TZP | FOX | AMK | GEN | CIP | TET | SXT | IPM | CHL |
|---|---|---|---|---|---|---|---|---|---|---|---|---|---|---|---|---|---|---|
| *E. coli* ST3274 | Pig | Nkolbisson | R | R | R | R | R | R | R | S | S | S | S | R | R | R | S | S |
| *E. coli* ST2705 | Pig | Odza | S | R | R | R | R | S | R | S | S | R | R | R | R | R | S | S |
| *E. coli* ST4389 | Pig | Afanoyoa | R | R | R | R | R | R | R | R | R | R | S | R | R | R | R | R |
| *E. coli* ST3716 | Environment | Nkolbisson | R | R | R | R | R | R | R | S | S | R | R | R | R | R | S | R |
| *E. coli* ST410 | Human | Odza | R | R | R | R | R | R | R | R | S | R | R | R | R | R | R | S |
| *E. coli* ST4684 | Human | Nkolbisson | S | R | R | R | R | S | R | S | S | S | R | R | R | S | R | R |
| *E. coli* ST3580 | Environment | Afanoyoa | R | R | R | R | R | S | R | S | S | S | S | R | R | R | S | S |
| *E. coli* ST3580 | Human | Afanoyoa | R | R | R | R | R | R | R | R | S | R | S | R | R | R | S | R |
| *K. quasipneumoniae* ST1535 | Pig | Etoudi | R | R | R | R | R | R | R | R | R | R | R | R | R | R | R | R |
| *K. quasipneumoniae* ST1535 | Human | Etoudi | R | R | R | R | R | R | R | S | S | R | S | S | S | S | S | S |

Abbreviations: AMP, ampicillin; AMX, amoxicillin; CFM, cefixime; CTX, cefotaxime; CAZ, ceftazidime; FEP, cefepime; ATM, aztreonam; TZP, piperacillin/tazobactam; FOX, cefoxitin; AMK, amikacin; GEN, gentamicin; CIP, ciprofloxacin; TET, tetracycline; SXT, sulfamethoxazole/trimethoprim; IPM, imipenem; CHL, chloramphenicol. R, resistant; S, susceptible.
